# Impaired spatial dynamic functional network connectivity and neurophysiological correlates in functional stroke mimics

**DOI:** 10.1101/2024.09.05.24313161

**Authors:** E Premi, V Cantoni, A Benussi, A Iraji, VD Calhoun, D Corbo, R Gasparotti, M Tinazzi, B Borroni, M Magoni

## Abstract

The present study investigated spatial dynamic functional network connectivity (dFNC) in patients with functional hemiparesis (i.e., functional stroke mimics, FSM). The aim of this work was to assess dynamic brain states, which represent distinct dFNC patterns that reoccur in time and across subjects. Resting-state fMRI data were collected from 15 patients with FSM (mean age=42.3±9.4, female=80%) and 52 age-matched healthy controls (HC, mean age=42.1±8.6, female=73%).

Each patient underwent a resting-state functional MRI scan for spatial dFNC evaluation and transcranial magnetic stimulation protocols for indirect assessment of GABAergic and glutamatergic transmission. We considered three dynamic brain networks, i.e., the somatomotor network (SMN), the default mode network (DMN) and the salience network (SN), each summarized into four distinct recurring spatial configurations. Compared to HC, patients with FSM showed significant decreased dwell time, e.g. the time each individual spends in each spatial state of each network, in state 2 of the SMN (HC vs. FSM, 13.5±27.1 *vs.* 1.9±4.1, *p*=0.034). Conversely, as compared to HC, FSM spent more time in state 1 of the DMN (10.8±14.9 *vs.* 27.3±38.9, *p*=0.033) and in state 3 of the SN (23.1±23.0 *vs.* 38.8±38.2, *p*=0.001). We found a significant correlation between the dwell time of impaired functional state of the SMN and measures of GABAergic neurotransmission (*r*=0.581, *p*=0.037). Specifically, longer impaired dwell time was associated with greater GABAergic inhibition. These findings demonstrate that FSM present altered functional brain network dynamics, which correlate with measures of GABAergic neurotransmission. Both dFNC and GABAergic neurotransmission may serve as potential targets for future intervention strategies.

## INTRODUCTION

Functional neurological disorders, including those mimicking acute strokes, also known as “functional stroke mimics” (FSM), present a significant challenge in clinical neurology due to their complex pathophysiology and the absence of identifiable structural brain lesions (1,2). These disorders, characterized by neurological symptoms such as hemiparesis without corresponding brain pathology, often lead to significant disability and healthcare utilization (3). Recent advances in neuroimaging and neurophysiology have provided new insights into the underlying mechanisms of these disorders, particularly through the study of functional brain connectivity and GABAergic neurotransmission (4). Investigations into the neurophysiological underpinnings of FSM have revealed valuable insights into motor cortex excitability and intracortical inhibition (5). A key finding is the significant increase in resting motor threshold (RMT) and short-interval intracortical inhibition (SICI) in the primary motor cortex contralateral to the affected limb in patients with FSM stroke mimics (6,7). This increase in SICI, which is not observed for intracortical facilitation (ICF), suggests an enhanced GABAergic inhibitory tone in the affected motor cortex compared to healthy controls (5).

Functional connectivity studies using resting-state functional MRI (rs-fMRI) have shown that FSM exhibits increased regional homogeneity (ReHo) in the left precentral gyrus, contralateral to the hemiparetic side, indicating abnormal local connectivity in motor-related areas (6). Concurrently, reduced connectivity between the right temporoparietal junction and bilateral sensorimotor regions suggests disrupted connectivity in regions associated with self-referential processing and sensory information integration (8). These alterations in functional connectivity highlight the complex interplay between motor execution and higher-order cognitive functions in FSM, supporting the hypothesis that FSM is underpinned by extensive network dysfunction rather than localized structural abnormalities, with multiple large-scale networks involved at the same time in the disease (9–11).

In this view, studying FSM using time-varying (dynamic) functional network connectivity (dFNC) represents a significant advancement in understanding the intricate neural mechanisms underlying these conditions (12). Unlike traditional static functional connectivity (sFNC) analyses, which provide an averaged view of brain network interactions over time, dFNC captures the moment-to-moment fluctuations in connectivity between brain regions (13). This dynamic approach is particularly valuable for detecting subtle perturbations in brain functioning that may not be evident in static measures (14). In particular, every brain network may have different configurations in space (i.e., “dissemination in space”) that can vary during the scan period (i.e., “dissemination in time”) (15–17). This spatial chronnectome approach leverages the spatial information of the variations of the spatial patterns of a given network to capture network-specific dynamics information (15–17). By examining how connectivity patterns change over time, researchers can uncover transient disruptions and abnormal network dynamics that contribute to the manifestation of FSM symptoms (18). This enhanced sensitivity to temporal variations in brain connectivity may offer a more comprehensive understanding of the neural dysfunctions in FSM, paving the way for more accurate diagnoses and targeted therapeutic interventions (19,20).

This approach may also shed light on the correlation between brain changes in FSM and already reported abnormalities of motor cortex excitability and short-interval intracortical inhibition (SICI), an indirect measure of GABAergic neurotransmission assessed with transcranial magnetic stimulation (TMS), in the primary motor cortex contralateral to the affected limb in patients with FSM (6,7,21). This change in SICI suggests an enhanced GABAergic inhibitory tone in the affected motor cortex compared to healthy controls (7).

These premises defined the objective of the present study, which aimed at evaluating changes in sFNC and dFNC in FSM, and their association with GABAergic and glutamatergic neurotransmission.

## METHODS

### Participants

Fifteen patients admitted at the Stroke Unit, ASST Spedali Civili Hospital, Brescia, Italy, for a suspected acute stroke and unremarkable brain structural imaging were recruited (8 right-side and 7 left-side hemiparesis). At 24 h follow-up, brain MRI scan with T1, T2-Fluid Attenuated Inversion Recovery (FLAIR) and Diffusion Weighted Images (DWI) sequences resulted in unremarkable and ruled out a cerebrovascular event or any other abnormality that could have possibly explained the symptomatology. Cervical spine MRI and EMG-ENG also resulted within the normal range.

At discharge (within 7 days after admission), global neurological examinations showed persistent hemiparesis, and a diagnosis of FSM was postulated. Patients eligible for inclusion were free from substance use disorder, bipolar disorder, schizophrenia, psychogenic non-epileptic seizure or any chronic or acute organic neurological disorder.

For the purpose of the present study, 52 age-matched healthy controls (HC) were recruited for comparison.

Each patient with FSM (within 3 months from the onset of symptoms) and HC underwent resting state functional MRI (rs-fMRI) to assess sFNC and dFNC, and TMS protocols to indirectly evaluate GABAergic and glutamatergic neurotransmission.

Informed consent was acquired from all participants in accordance with the Declaration of Helsinki. The local ethics committee of the Brescia Hospital approved the present study (05.19.2015, #NP1965). The present study complies with the STARD guidelines (https://www.equator-network.org/reporting-guidelines/stard/).

### MRI acquisition and preprocessing

Each subject underwent an MRI scan and brain images were collected using two different Siemens (Siemens, Erlangen, Germany) scanners (Siemens Avanto 1.5T: 5 FSM and 23 HC; Siemens Skyra 3T: 10 FSM and 29 HC). T2-weighted echo planar imaging (EPI) sequences sensitized to blood oxygenation level-dependent (BOLD) contrast for rs-fMRI were considered (Siemens Avanto 1.5T: 200 timepoints, repetition time [TR] = 2500 ms, echo time [TE] = 50 ms; Siemens Skyra 3T: 200 timepoints, repetition time [TR] = 2500 ms, echo time [TE] = 30 ms), as previously published (22). During scanning, subjects were asked to keep their eyes closed, not to think of anything in particular, and not to fall asleep. Functional data were preprocessed using the toolbox for Data Processing & Analysis for Brain Imaging (DPABI, http://rfmri.org/dpabi) (23) based on Statistical Parametric Mapping (SPM12, https://www.fil.ion.ucl.ac.uk/spm/) software, as previously reported (17,22). For each subject, the first 5 volumes of the MRI series were discharged to account for magnetization equilibration. The remaining 195 volumes underwent slice-timing correction and were realigned to the first volume. Any subject with a maximum displacement in any direction larger than 2.5 mm, or a maximum rotation (x,y,z) larger than 2.5°, was excluded. Moreover, we considered framewise displacement (FD) (24) as a nuisance variable accounting for head motion during MRI scanning. Data were subsequently normalized to the EPI unified segmentation (25) in Montreal Neurological Institute coordinates derived from SPM12 software and resampled to 3×3×3 cubic voxels. Spatial smoothing with an isotropic Gaussian kernel with the full width at half-maximum (FWHM) of 6 mm was applied, followed pre-processing pipeline previously adopted for spatial chronnectome analysis (15,17).

### Functional networks decomposition

MRI data were processed using the GIFT (GIFT toolbox, http://trendscenter.org/software/gift)(26), and a spatially constrained multivariate objective optimization ICA with reference (MOO-ICAR) (27,28) was used to obtain spatial maps from a set of selected large-scale networks (Iraji et al., 2019). In the present study, we considered the somato-motor network (SMN), the default mode network (DMN) and the salience network (SN) (15). Spatial maps are used as reference templates to calculate functional networks for each subject by maximizing independence in the context of the spatial constraint, as already described (15,29).

### Static functional network connectivity (sFNC) analysis

Individual networks were estimated with MOO-ICAR preprocessing (15). Back-reconstruction step considered the estimation of subject-specific networks and their related time courses based on the selected 3 networks (SMN, DMN, and SN) (15,30). Statistical analysis was then performed using SPM12, as follows: *a)* between-group comparison (FSM vs HC), considering scanners site and FD Powers as nuisance variables (*p*<0.001, uncorrected for multiple comparisons); *b)* multiple regression to assess the relationship between imaging variables and neurophysiological variables, covarying for scanner type and FD Power (*p*<0.001, uncorrected for multiple comparisons).

### Dynamic functional network connectivity (dFNC) analysis

The dFNC analysis was achieved using the dFNC toolbox implemented in GIFT (GIFT toolbox, http://trendscenter.org/software/gift) (26). Sliding window length or number of clusters were chosen according to previous literature data on dynamic connectivity (15,16,26). The single time courses were detrended (to remove baseline drifts from the scanners and/or physiological pulsations), orthogonalized with respect to 12-motion parameters, despiked (replacement of outlier time points with 3^rd^ order spline fitting to clean neighboring points) and filtered using a 5^th^ order Butterworth filter (0.01–0.15 Hz). For each considered brain network, the temporal coupling between a specific brain network and every voxel of the brain was calculated using the sliding-window correlation approach resulting in one dynamic coupling map (dCM) per window. This procedure takes all the potential associations into account and fully captures the relationship between each voxel and the brain network (for example, if a given voxel is highly correlated with two networks, correlation analysis allows the detection of both of these associations). We used the tapered window obtained by convolving a rectangle (width=30 TRs) with a Gaussian (σ=3 TRs) and the sliding step size of one TR (15,16).

*k*-means clustering was applied to summarize the dCMs of each brain network into a set of spatial states, which allows us to investigate the dynamic properties of the brain network via temporal variations of these distinct spatial states. The number of spatial states was set to 4, in line with Iraji et al. (15). Using temporal profiles of the spatial states, the mean dwell time (DT), that is the average of the amount of time that subjects stay in a given state once entering that state, was calculated for each network, as state-level dynamic index to summarize dynamic properties of each network.

### Transcranial magnetic stimulation (TMS) protocols

A TMS figure-of-eight coil (each loop diameter 70 mM – D702 coil) connected to a monophasic Magstim Bistim2 system (Magstim Company, Oxford, United Kingdom) was employed for all TMS paradigms, as previously reported (31). Motor evoked potentials (MEPs) were recorded from the right and left first dorsal interosseous (FDI) muscles through surface Ag/AgCl electrodes placed in a belly-tendon montage and acquired using a Biopac MP-150 electromyograph (BIOPAC Systems Inc., Santa Barbara, CA, United States). Responses were amplified and filtered at 20 Hz and 2 kHz with a sampling rate of 5 kHz and recorded on a personal computer for offline elaboration (AcqKnowledge 4.1, BIOPAC Systems Inc., Santa Barbara, CA, United States).

Resting motor threshold (RMT) was determined contralateral to hemiparetic side as the minimum intensity of the stimulator required to elicit motor evoked potentials (MEPs) with a 50 mV amplitude in 50% of 10 consecutive trails, recorded from the right or left first dorsal interosseous muscles during full relaxation (32). Moreover, MEP latencies were measured on both sides at an intensity of 120% RMT.

We considered short interval intracortical inhibition (SICI), an indirect marker of GABAergic neurotransmission, and intracortical facilitation (ICF), an indirect marker of glutamatergic neurotransmission, which were studied using a paired-pulse technique, employing a conditioning-test design. For all paradigms, the test stimulus (TS) was adjusted to evoke a MEP of 1 mv amplitude in the right and left FDI muscles (31).

The conditioning stimulus (CS) was adjusted at 70% of the RMT, employing multiple interstimulus intervals (ISIs), including 1, 2, 3 ms for SICI and 7, 10, 15 ms for ICF (33,34). For each ISI and for each protocol, ten different paired CS-TS stimuli and fourteen control TS stimuli were delivered in all participants in a pseudo-randomized sequence, with an inter trial interval of 5 s (10%). The conditioned MEP amplitude, evoked after delivering a paired CS-TS stimulus, was expressed as percentage of the average control MEP amplitude.

### Statistical analyses

Comparisons of demographic and clinical characteristics were assessed by Student’s t-test for continuous variables and χ^2^ test for categorical variables. We compared dFNC indexes between FSM vs HC with univariate general linear model (GLM) analyses, considering scanner type and framewise displacement (FD Power) as nuisance variables. TMS measures were compared using a two-way repeated measures ANOVA. If a significant main effect was obtained, group differences were examined with post hoc tests (Bonferroni correction for multiple comparisons). Partial correlation analyses were run to compare dFNC measures, neurophysiological parameters and clinical variables considering scanner type and FD Power as covariates.

Statistical analyses were performed using IBM SPSS Statistics 29.0 (Chicago, USA) and the statistical significance level was set at *p*<0.05.

### Data availability

All study data, including study design, protocol, statistical analysis plan, and results are available from the corresponding author, upon reasonable request.

## RESULTS

### Participants

Demographic and clinical characteristics of patients with FSM and age-matched HC are reported in **Table 1**.

**Table 1.**
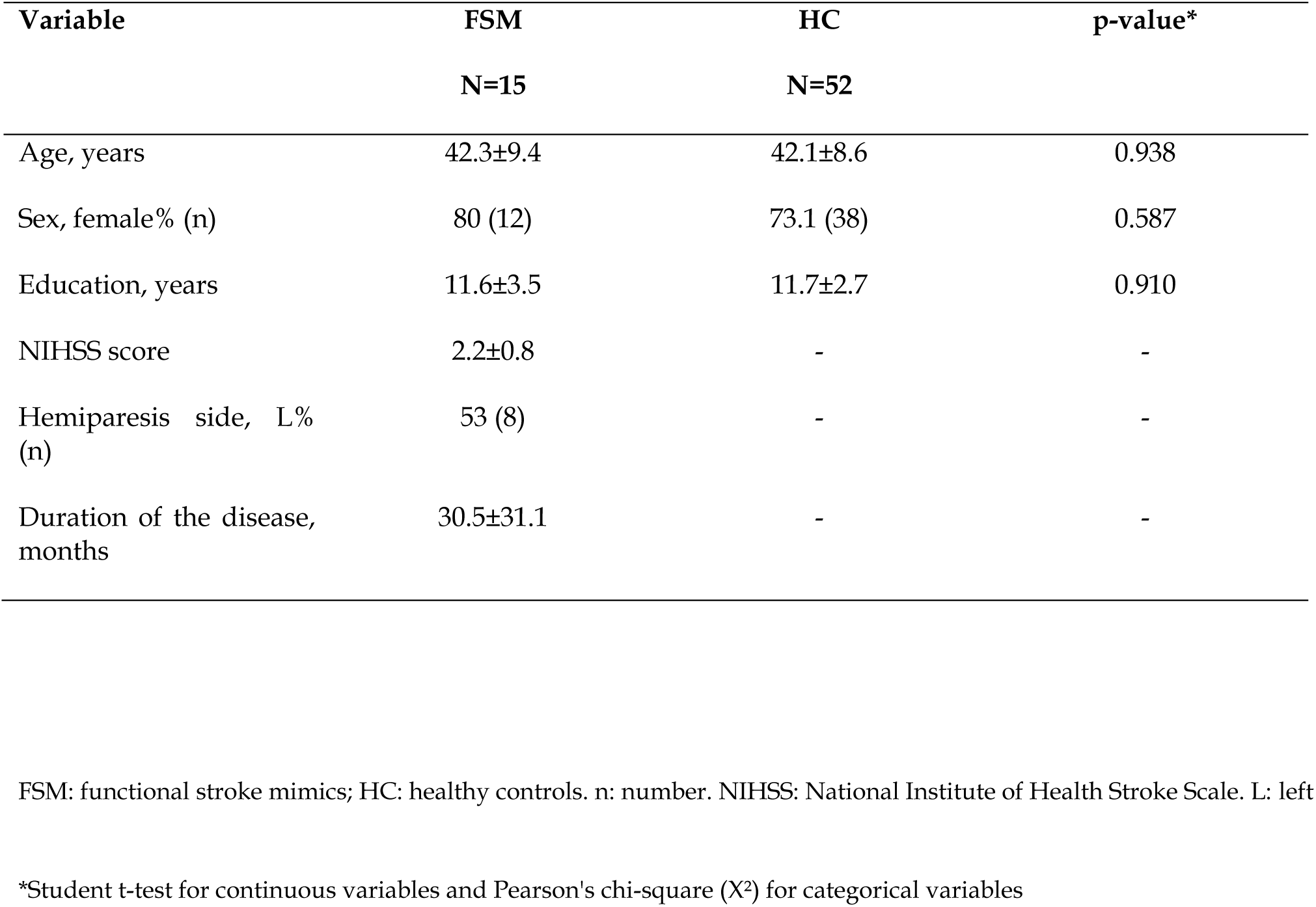
Demographic and clinical characteristics of patients with FSM and HC.

We estimated the sFNC and dFNC of the SMN, the DMN and the SN in the overall sample of subjects. Network patterns assessed for sFNC resembled those already reported in the literature (see **Figure 1**); dFNC analyses demonstrated different patterns across four states for each considered network, as already demonstrated (15).

**Figure 1.**
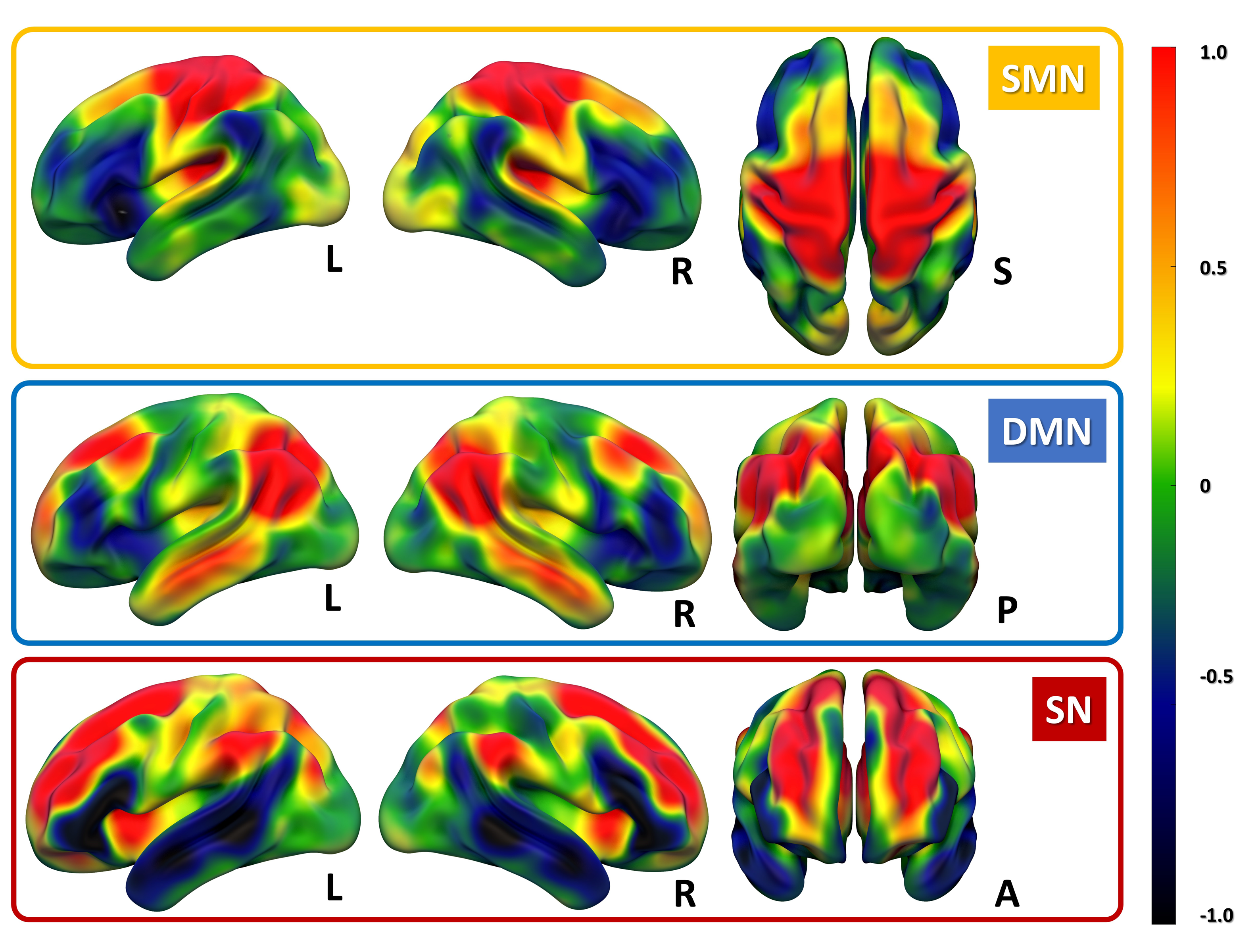
The somatomotor network (SMN), the default mode network (DMN) and the salience network (SN) patterns considered for static Functional Network Connectivity analyses. Hot and cold colors represent positive and negative correlations among different brain regions, respectively. Maps of each network are displayed on a standardized 3D T1 MRI template (https://www.nitrc.org/projects/surfice/). L: left, R: right, S: superior, P: posterior, A: anterior.

The four states of the SMN were defined by signal in sensorimotor structures, with slight variations across states. In particular, state 2 showed a less pronounced involvement of sensorimotor structures, while state 4 showed an anticorrelation with parietal regions (see **Figure 2**).

**Figure 2.**
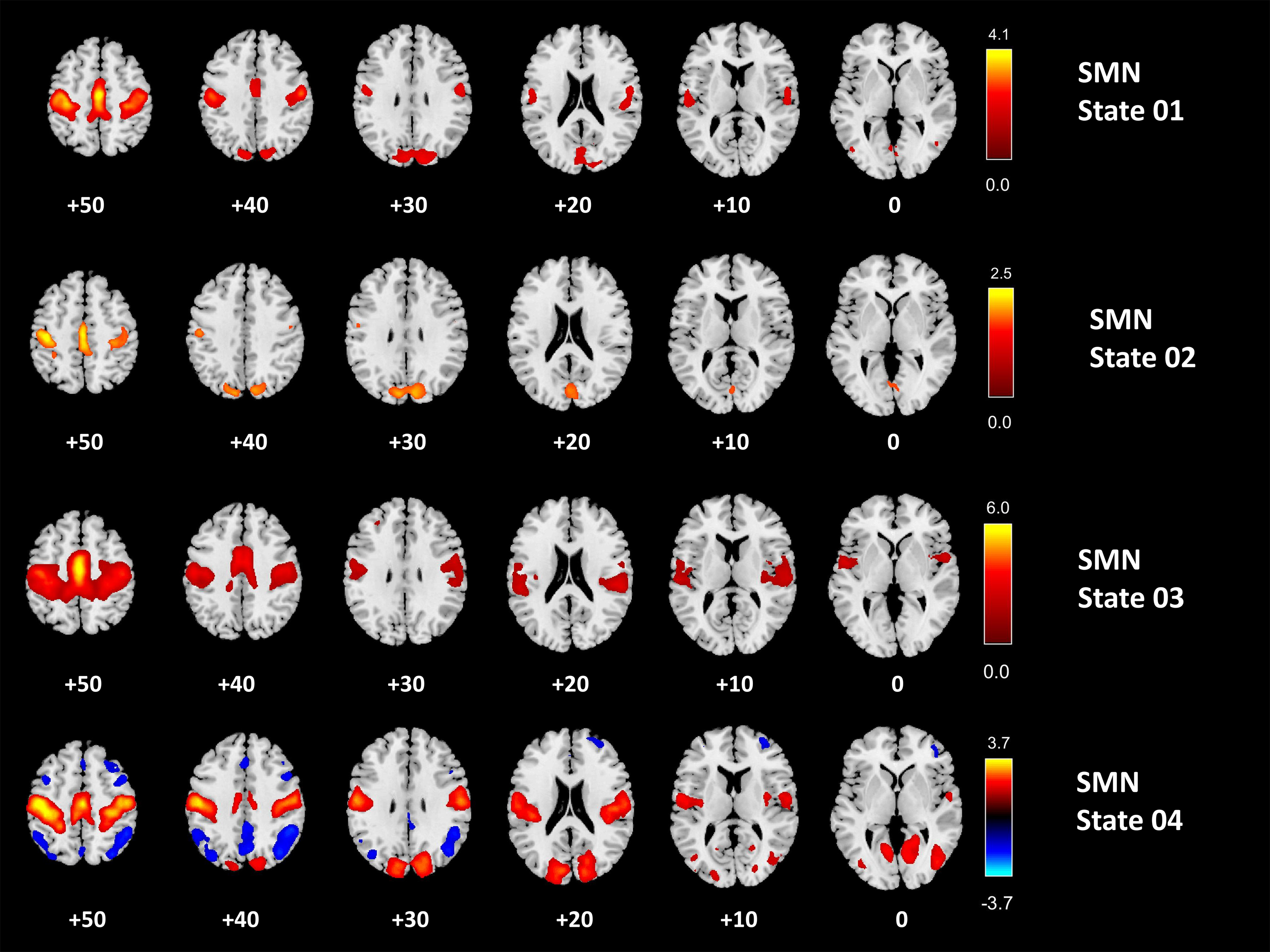
Spatial states of the somatomotor network (SMN). Hot and cold colors represent positive and negative associations to the SMN, respectively. Maps of each spatial states are displayed on a standardized axial T1 MRI template, z-axis coordinates are reported under each slice.

The four spatial states of the DMN were characterized by the posterior cingulate cortex and bilateral parietal hubs, in association with a small cluster in the anterior cingulate (see **Figure 3**). This pattern was evident in states 1, 3, and 4, with state 2 demonstrating an involvement more restricted to the posterior regions.

**Figure 3.**
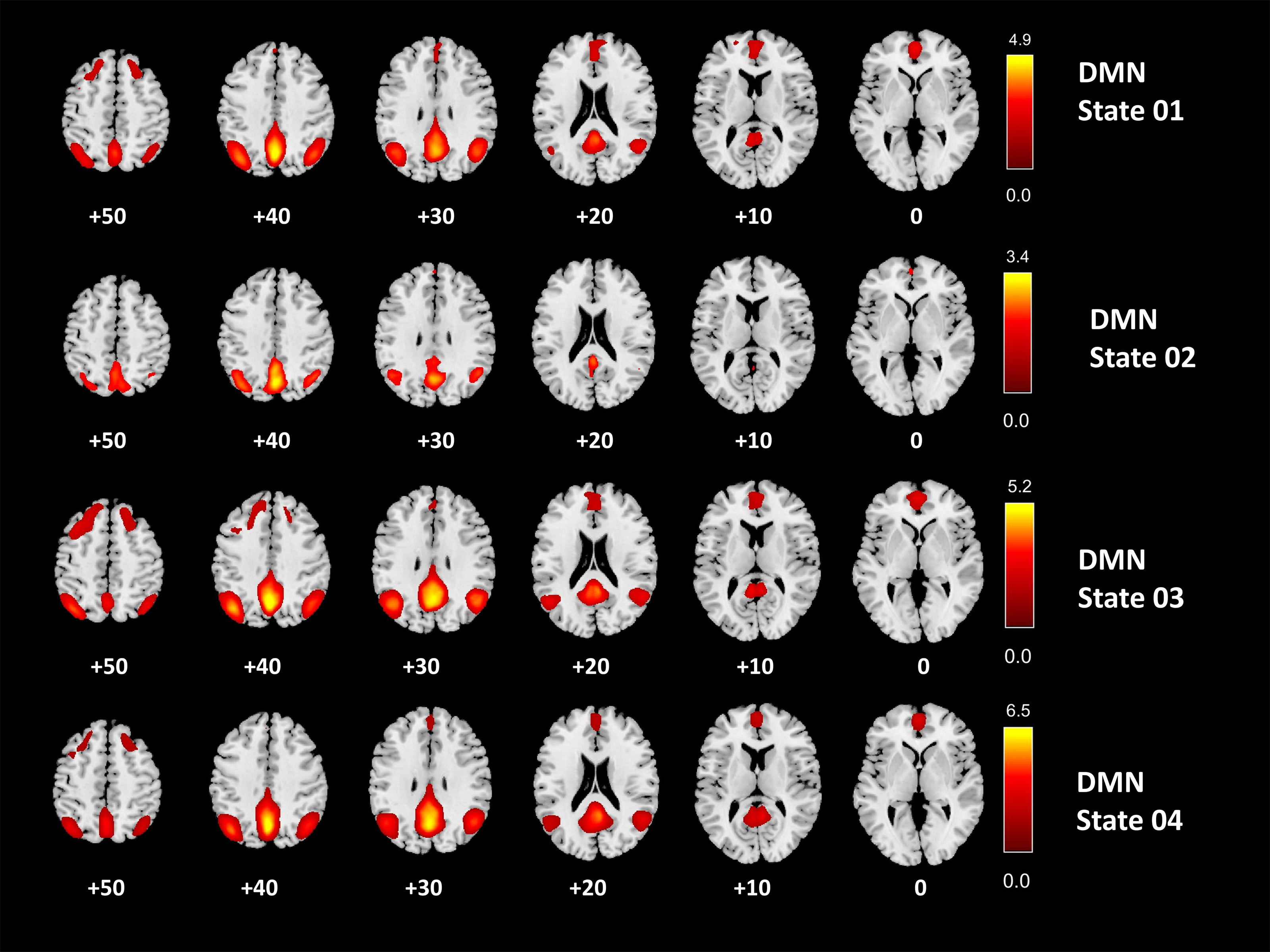
Spatial states of the default mode network (DMN). Hot and cold colors represent positive and negative associations to the DMN, respectively. Maps of each spatial states are displayed on a standardized axial T1 MRI template, z-axis coordinates are reported under each slice.

The four spatial states of the SN were characterized by a frontal hub, with a more pronounced signal in the insula region, bilaterally. State 1 and state 4 showed an anticorrelation with parietal and temporal regions, while state 3 was defined by an anticorrelation with frontal regions (see **Figure 4**).

**Figure 4.**
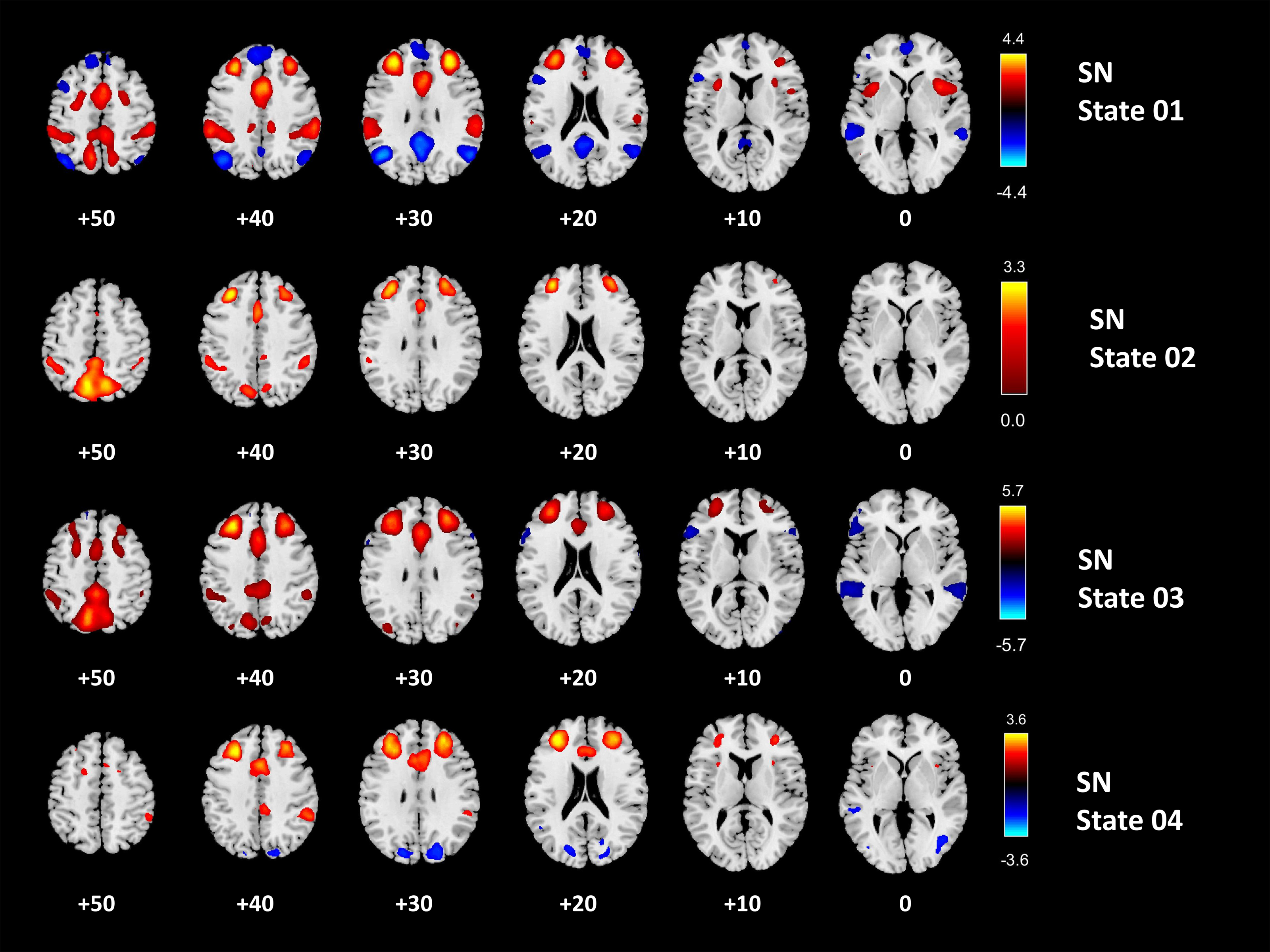
Spatial states of the salience network (SN). Hot and cold colors represent positive and negative associations to the SN, respectively. Maps of each spatial states are displayed on a standardized axial T1 MRI template, z-axis coordinates are reported under each slice.

As already demonstrated, when we considered TMS parameters, patients with FSM showed impaired SICI, an indirect measure of GABAergic neurotransmission (p < 0.05 FSM vs. HC using one-way ANOVA, post hoc tests with Bonferroni correction for multiple comparisons), while no significant changes were reported in regard of ICF, an indirect measure of glutamatergic neurotransmission (see **Supplementary Figure 1**).

### Analysis of sFNC

When we considered spatial maps of sFNC, no significant differences were observed between FSM and HC across the three networks considered at the pre-established threshold. Moreover, no significant correlations were found between sFNC and TMS indexes (SICI or ICF).

### Analysis of dFNC

Considering dFNC, when we compared dwell time (DT), e.g., the amount of time that subjects stay in a given state, we found significant differences between FSM and HC. FSM participants spent significantly less time in state 2 of the SMN, the state with less pronounced involvement of sensorimotor structures, compared to HC (HC *vs.* FSM, 13.5±27.1 *vs.* 1.9±4.1, *p*=0.034), (see **Table 2**). Conversely, FSM participants spent significantly more time in state 1 of the DMN (10.8±14.9 vs. 27.3±38.9, *p*=0.033) and in state 3 of SN (23.1±23.0 *vs.* 38.8±38.2, *p*=0.001) compared to HC (see **Table 2**).

**Table 2.**
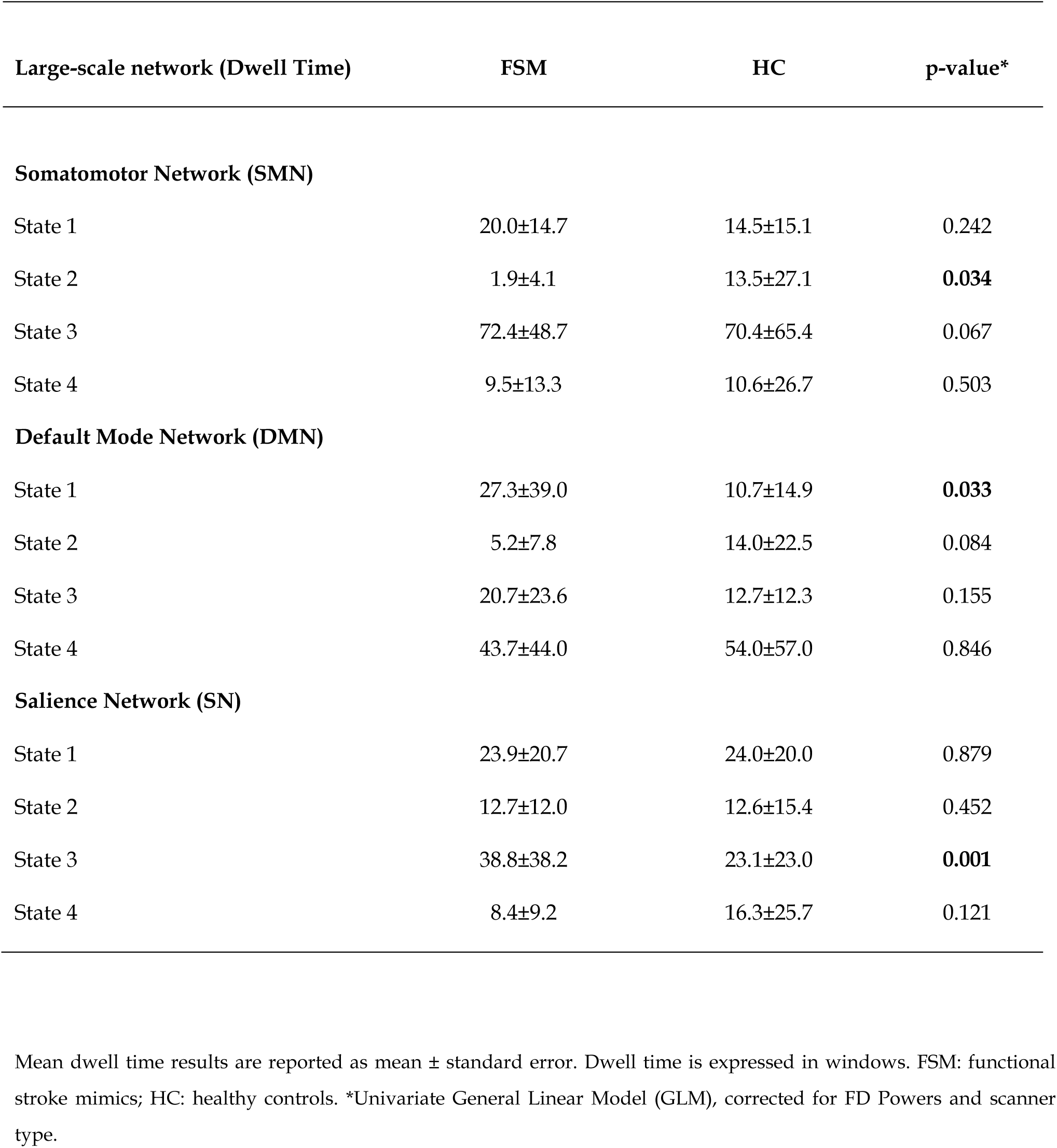
Mean dwell time (in each spatial state according to considered dynamic functional networks) in patients with FSM and HC.

### Correlations of dFNC measures and neurophysiological variables

When we evaluated the association between DT and TMS parameters, we found that DT of state 2 in the SMN positively correlated with SICI (*r*=0.581, *p*=0.037). The shorter the DT in state 2 (where participants with FSM spent less time compared to HC), the lower the SICI (indicating higher GABAergic inhibition). Conversely, the DT of state 3 in the SMN, which is characterized by a more pronounced involvement of sensorimotor structures, is inversely correlated with SICI (*r*=-0.668, *p*=0.013). Additionally, the DT of state 4 in the SN, which is anticorrelated with bilateral parietal regions, positively correlated with SICI (*r*=0.702, *p*=0.007).

### Correlations of clinical variables with neurophysiological variables and dFNC measures

We found a significant correlation between NIH Stroke Scale scores and SICI (r= 0.676, *p*=0.011), as well as with dFNC measures, including the DT of state 2 in the SMN (r= 0.715, *p*=0.006), the DT of state 3 in the SMN (r= -0.781, *p*=0.002), and the DT of state 4 in the SN (r= 0.607, *p*=0.028). Moreover, when considering the duration of symptoms, a similar pattern of association with dFNC measures was found (SMN state 2 DT: *r*=0.810, *p*=0.001; SMN state 3 DT: *r*=-0.672, *p*=0.012; SN state 4: *r*=0.595, *p*=0.032).

## DISCUSSION

FSM constitutes a significant proportion of cases in acute stroke services and place a considerable burden on healthcare resources. Despite recent advances in neurobiological understanding of FSM, the underlying mechanisms remain to be fully elucidated.

Previous functional MRI studies have provided converging evidence that distinctive neural network changes are involved in the pathophysiology of functional movement disorders (35–37). So far, functional connectivity studies have used static functional connectivity analyses, which are applied under the assumption that connectivity patterns of spatially separated brain regions remain constant over time. However, the connectivity patterns between brain regions fluctuate over time, and these patterns tend to return across time (12). This results in a number of identifiable spatial states, that may recruit distinct neuroanatomical brain regions, for each considered functional brain network (i.e., the DMN).

In this study, we evaluated both sFNC and dFNC in FSM, considering networks of potential interest, such as the SMN, DMN, and SN, each represented by four distinct recurring spatial configurations. While no significant differences were observed in sFNC, the specific changes in dFNC provided further insights into the neuronal mechanisms underlying the pathophysiology of FSM.

Patients with FSM spent significantly less time in SMN state characterized by lower activity in the sensorimotor structures compared to other SMN states. Additionally, patients spent more time in one DMN state, which is characterized by the typical activation pattern of the posterior cingulate cortex and bilateral parietal hubs, along with a small cluster in the anterior cingulate. They also spent more time in one SN state, which is characterized by its anticorrelation with frontal regions, compared to other SN states. Furthermore, disease severity, as measured by the NIH Stroke Scale, showed a significant correlation with neurophysiological parameters (SICI) and with dFNC measures. Symptom duration was also significantly correlated with dFNC measures.

Our results support the concept that the pathophysiology of FSM involves multiple brain networks, including those related to motor control (i.e., SMN), limbic/emotion regulation (i.e., SN), and self-agency and multimodal integration (i.e., DMN) circuits. Taken together, these findings suggest alterations in the dynamic neural architecture underlying pathological afferent and efferent higher-order motor processes in FSM (11). Interestingly, these findings were detectable only through the dynamic approach, resonating with Charcot’s idea of a “dynamic lesion” that eludes our “present means of investigation” as the neural basis of FSM (38–40).

Another key finding of this study was that abnormalities in dFNC were significantly correlated with SICI, an indirect measure of GABAergic neurotransmission (7).

SICI is thought to reflect short-lasting post-synaptic inhibition, partially mediated by GABAergic interneurons, which modulate corticospinal output activity (41). Impairment in SICI could indicate an increase of inhibitory intracortical circuits, affecting corticospinal excitability and potentially leading to a decrease in voluntary movements. In our study, we observed that the greater the changes in the time spent in the SMN state with reduced signal in sensorimotor areas, the greater the GABAergic inhibition. Similarly, we found a significant inverse correlation between the SMN state with increased signal in sensorimotor areas and GABAergic inhibition. Although preliminary, these findings suggest a potential link between imaging abnormalities in the brain chronnectome and changes in neuronal circuits related to GABAergic inhibition.

We acknowledge several limitations in our study. First, there relatively small sample size may have increased the risk of false-negative findings. However, compared to other studies on functional movement disorders, we recruited a well-characterized and homogeneous group of participants with isolate long-lasting flaccid functional paralysis, studied with a multimodal approach (dFNC and TMS). Nonetheless, it is important to note that our findings did not survive correction for multiple comparisons, which may weaken the strength of our conclusions. Despite this, the present findings should be viewed as an interesting starting point for the multimodal study of FSM.

In conclusion, our study demonstrates altered brain network dynamics in FSM, supporting the concept of an imbalance across brain networks involved in motor control and sense of agency. Future studies should exploit the chronnectome framework across the spectrum of functional movement disorders to further extend and consolidate the evidence herein described.

---

**Supplementary Figure 1. Neurophysiological parameters in patients with functional stroke mimics (FSM).** Short-interval intracortical inhibition (SICI) at ISI 1, 2, 3 ms and intracortical facilitation (ICF) at ISI 7, 10, 15 ms. Data are represented as a ratio to the unconditioned MEP amplitude; error bars represent standard errors. MEP = motor evoked potential; ISI = inter stimulus interval. Dotted line indicates mean values in age-matched healthy control group (n=10, age, 44.0±6,5 years; gender, female% (n)=40% (4)). *p < 0.05 FSM vs. HC using one-way ANOVA (post hoc tests with Bonferroni correction for multiple comparisons).

